# How do autistic adults and their families experience, understand, and manage gastrointestinal (GI) symptoms in daily life? *A qualitative manuscript from the “GI Wanna Talk About Autism” Study*

**DOI:** 10.64898/2026.01.13.26343894

**Authors:** Katherine Almendinger, Calliope Holingue, Margaret Faucett, Faith DiJulia, Kelsie Daley, Zachary Williams, Susan Brasher, Joseph J. Gallo, Elizabeth Pellicano

**Author notes:** Authors contributed equally (Katherine Almendinger, Calliope Holingue, Margaret Faucett).

## Abstract

**Background:** Gastrointestinal (GI) symptoms are prevalent, persistent, and frequently disabling among autistic individuals. Existing research has focused predominantly on children, with comparatively little attention to GI experiences in adulthood. Qualitative studies in pediatric populations document substantial unmet GI-related healthcare needs, including negative healthcare encounters and limited access to autism-informed services.

**Methods:** Using a community-based participatory research (CBPR) framework, we conducted qualitative interviews with 26 participants (21 autistic adults, 5 parents reporting on behalf of an adult autistic child), of varying race, sexual orientation, genders, socioeconomic and educational statuses, and ages. Interviews were conducted on Zoom, ranging from 22–110 minutes long, exploring the physical, emotional, and functional impacts of GI symptoms; how these experiences relate to autism; barriers to treatment; and participants’ needs and priorities for improving GI health care (priorities reported elsewhere). We conducted a reflexive thematic analysis following Braun and Clarke, using an interpretivist-constructivist epistemological stance. Coding and theme development were inductive and data-driven. Themes were refined collaboratively through repeated engagement with the data, analytic memoing, and discussion of areas of interpretive uncertainty until shared meaning and coherent thematic structure were achieved. Once the codebook and thematic structure were finalized, all transcripts were systematically coded for analysis.

**Results:** Participants described gastrointestinal symptoms as chronic, unpredictable, and highly consequential, shaping physical functioning, emotional wellbeing, daily routines, autonomy, and social participation. Symptoms were understood as arising from interacting biological, sensory, emotional, and contextual factors, with triggers often difficult to identify or anticipate. Experiences with healthcare were frequently characterized by dismissal, communication barriers, system complexity, and prior trauma, contributing to delayed or avoided care and heightened distress. In response, autistic adults and caregivers relied on individualized, trial-and-error management strategies – including avoidance of triggers, routine and environmental planning, dietary and pharmacologic approaches, and sensory or emotional regulation – alongside social support and peer communities to cope with persistent uncertainty and limited clinical guidance.

**Conclusion:** GI symptoms in autistic adults frequently have dramatic negative impacts on everyday life, reducing both quality of life and restricting the ability to fully engage in society and desired activities. Despite the clear magnitude of impact, knowledge and support are lacking and management remains difficult, confusing, and often unsuccessful. Improving care will require multi-layered, neurodiversity-informed approaches that recognize autistic adults as central knowledge-holders and active partners in research and clinical decision-making.

## Introduction

Autistic individuals experience higher rates of both psychiatric and medical conditions compared to non-autistic individuals^1,2^ – anxiety and mood disorders,^2^ obesity, hypertension, diabetes,^1^ epilepsy, sleep disorders, gastrointestinal (GI) disorders,^2^ – which impact many aspects of everyday life^6,7^ and can negatively shape long-term wellbeing. In this study, we focus specifically on GI symptoms among autistic adults.

There is remarkably limited research on GI issues in autistic adults, despite most autistic people being adults^25^. Most literature focuses on children, though, even in children, prevalence estimates range widely due to the heterogeneity of autism and differences in measures used across studies.^4^ The literature suggests anywhere from 9%–91% of autistic children experience GI symptoms like diarrhea, constipation, and abdominal pain.^3^ Commonly reported GI symptoms include constipation, abdominal pain, reflux, and others.^5^

Health research in autistic populations has traditionally focused on discrete medical issues examined through quantitative methods. Quantitative approaches generally focus on “when,” “what,” or “where” questions addressed by collecting pre-defined data points from large populations through indirect means. In recent years, however, qualitative research has gained recognition for its ability to provide richer and more in_-_depth insights into lived experiences.^8^ A key advantage of qualitative approaches is their usefulness in contexts where variables cannot be tightly controlled, which is often the case in health research and is the case in this study as well.^9^ Qualitative research generally seeks to answer “how” and “why” questions by engaging individuals or small groups of people through more personal means. Groups are typically united by one or more common characteristic, such as a shared health condition,^8^ and tools such as interviews and free-form survey responses are employed to understand the meaning behind individuals’ experiences.^10^ This is particularly important in the case of GI-related symptoms as we know exceedingly little about the phenomenological experiences of symptoms for autistic people or how these symptoms affect a person’s sense of self, body, and world.

This study sought to address this gap by conducting semi-structured qualitative interviews of autistic adults with co-occurring GI issues to understand how autistic adults and their families experience, understand, and manage GI symptoms in daily life.

## Methods

Institutional Review Board Approval was granted by Johns Hopkins University: JHM IRB 00316729.

### Study Approach

We used a community-based participatory research (CBPR) approach, increasingly being used in autism research, to enhance the relevance, applicability, impact, and equity of scientific findings.^11^ The participatory research approach is centered around engaging the subjects of research and their communities, who normally would have been excluded from participating, in co-constructing research about or for those populations or communities.^26^

### Community Advisory Board Assembly and Involvement

At project outset, we assembled a Community Advisory Board (CAB) to serve as active research team members. CAB members were recruited through a variety of channels, including existing professional and personal connections, referrals from colleagues, outreach via social media, etc. The CAB provided input on all study activities and decisions, guiding our work to ensure accurate reflection of community priorities and lived experiences, but did not interact with participants.

The CAB, comprised autistic and non-autistic adults with diverse professional and lived experiences, included 24 members: six autistic adults, six parents or siblings of autistic adults, ten healthcare professionals, and two autism researchers. The CAB was actively involved in multiple aspects of the study, including reviewing all participant-facing materials (consent forms, questionnaires, and interview guides), providing feedback to improve our content, particularly the clarity and accessibility of our materials.

We prioritized accessible and flexible communication throughout the project. Both CH and MF were available via email to the entire CAB for regular communication and asynchronous feedback about the project. The full CAB met four times via Zoom, with closed captions enabled and transcripts recorded, to discuss study materials, qualitative results, and the creation of recommendations. Meeting materials were shared in advance, and written summaries were distributed afterward. Members could participate via microphone or chat, and one-on-one meetings were offered as needed.

Communication occurred frequently by email, either with the full group, subgroups, or individuals, and the team used Slack to support ongoing discussion. The lead researchers (CH, MF) were responsible for ensuring that all study activities and communications were as accessible as possible for the board. Although the two lead researchers retained final decision-making authority, they sought input from the CAB at every stage of the study and incorporated suggestions into study procedures.

### Study Team

The final study team included 30 individuals: the 25-member Community Board (one of whom participated in data analysis, FD), four participant-facing interviewing researchers [two autistic (MF, KA) and two non-autistic (CH, KD); includes the two co-principal investigators (CH, MF)], and a senior mentor with extensive experience conducting qualitative autism research using a CBPR approach (EP).

### Participant Recruitment

Recruitment occurred through investigator networks, autism-related listservs, and outreach by our funders. A study flyer was distributed via email, social media, community groups, and autism-related newsletters. The flyer included a link to a screening survey used to verify eligibility and identify individuals for qualitative interviews. Eligibility criteria were: 1) being 18 years of age or older, 2) self-identifying as being autistic (no formal diagnosis documentation was required, though many had been formally diagnosed), and 3) self-reporting a history of GI symptoms. Both autistic adults and parents/carers of autistic adults were eligible; however, we prioritized interviewing a larger number of autistic adults. The survey also collected demographic and clinical information, enabling us to purposively sample from a diverse range of participants (see Table 1 for final participant characteristics). Participants were paid $10/hour of interviewing, usually $10/interview.

**Table 1.** Characteristics of Study Participants Interviewed for Qualitative Analysis.

| **Demographics** | **Total n = 26** |
| --- | --- |
| **Age:** | **<u>Average Years (SD)</u>** |
| Age of autistic adults | 30 ($\pm 8$) |
| Age at autism diagnosis | 18 ($\pm 12$) |
| **Gender:** | **<u>n (%)</u>** |
| Man | 11 (42) |
| Nonbinary or Gender non-conforming | 4 (15) |
| Woman | 11 (42) |
| **Race:** |  |
| Other Race | 2 (8.3) |
| White | 22 (92) |
| **Education and Employment** | **<u>n (%)</u>** |
| Did not receive high school diploma/GED | 3 (12) |
| Received high school diploma/GED | 2 (7.7) |
| Some college, no degree | 3 (12) |
| Associate Degree | 3 (12) |
| Bachelor Degree | 8 (31) |
| Graduate degree | 7 (27) |
| Employed | 16 (62) |
| **Income:** |  |
| <\$35k USD | 8 (32) |
| \$35-50k USD | 3 (12) |
| \$50-75k USD | 5 (20) |
| \$75k+ USD | 9 (36) |
| **Living Situation** | **<u>n (%)</u>** |
| Live alone | 3 (12) |
| Live with one or more roommates | 4 (15) |
| Live with parents or other relatives | 10 (38) |
| Live with spouse/partner | 7 (27) |
| Other | 2 (7.7) |
| **Preferred Language** | **<u>n (%)</u>** |
| Want to be referred to: |  |
| As a person on the autism spectrum | 3 (12) |
| As a person with autism | 6 (23) |
| As an autistic person | 14 (54) |
| Other | 3 (12) |
| **GI History** | **<u>n (%)</u>** |
| GI symptoms in past 3 months | 25 (96) |
| Length of GI symptoms in past 3 months |  |
| Past 3 months only | 1 (4.0) |
| 1-5 years | 3 (12) |
| More than 5 years | 21 (84) |
| Ever had GI symptoms for 3+ months |  |
| Yes | 26 (100) |
| Have a GI Diagnosis | 15 (58) |
| GED=general educational degree; GI=gastrointestinal; SD=standard deviation; USD=United States Dollars. |  |

**Table 2.**
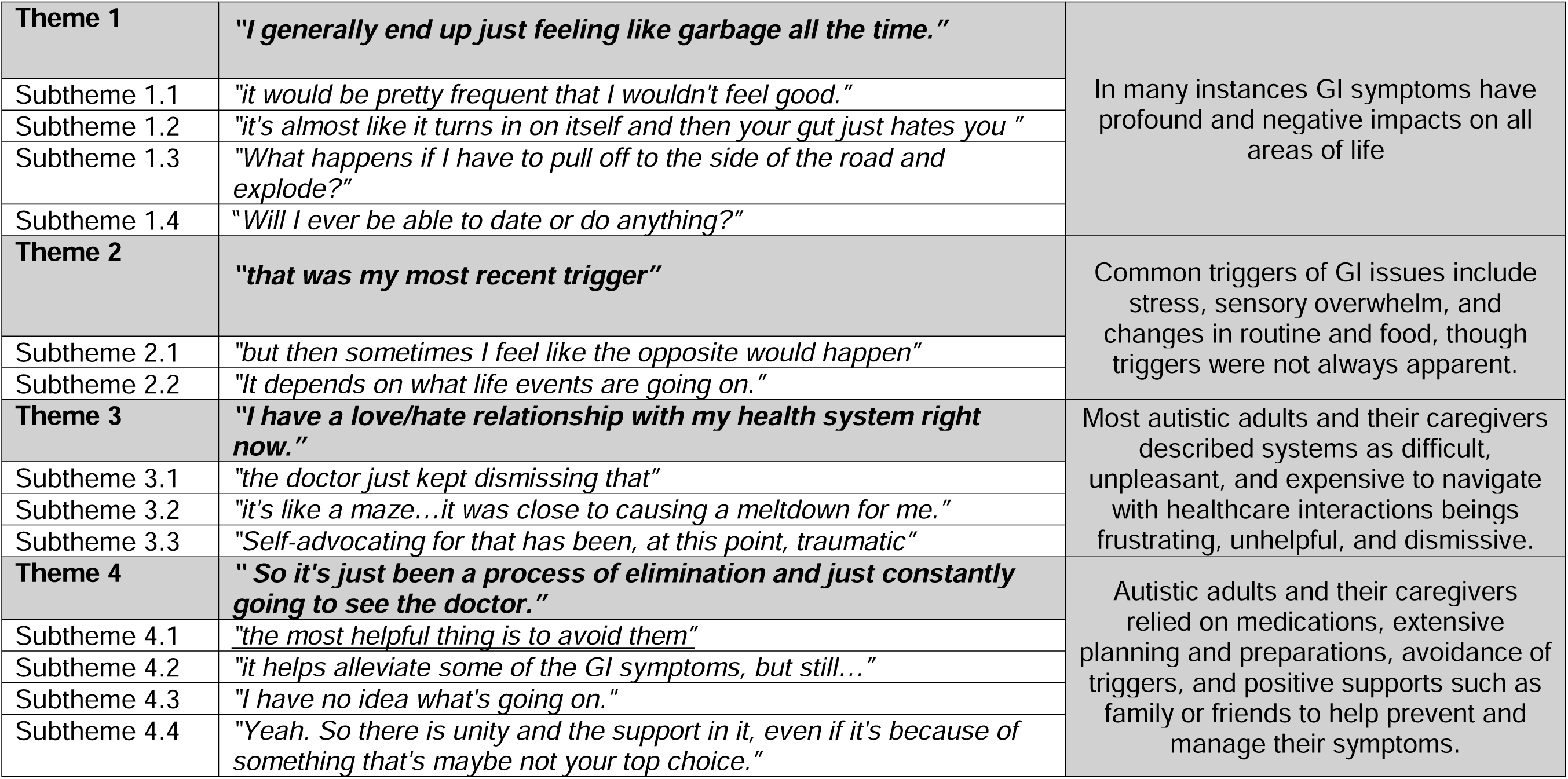
Key Qualitative Themes.

### Participants

Twenty-six participants took part in the study. Most (n = 19, 73%) were autistic adults with a professional diagnosis, two (8%) were adults without a professional diagnosis, but who self-identified as autistic, and five (19%) were parents reporting on behalf of their adult autistic child (Table 1).

Autistic participants were on average, 30 years old (SD ± 8 years), with autism diagnoses typically received around age 18 (range = 2–42 years). Most participants identified as White (n = 22, 92%) and there was an even split between men and women, with four participants selecting nonbinary or gender nonconforming (Table 1).

Nearly all participants (n = 25; 96%) reported GI symptoms in the past three months, with 22 (84%) experiencing a duration of more than five years and 100% reporting a duration of at least three months. Of those participating, the majority reported being diagnosed with Irritable Bowel Syndrome (n = 10, 38%), the second most common diagnosis was Gastroesophageal Reflux Disease (n = 6, 23%), followed by Chronic Constipation (n = 4, 15%).

### Semi-structured interviews

The interview focused on understanding how autistic adults experience and manage GI symptoms. It explored the physical, emotional, and functional impacts of GI symptoms; how these experiences relate to autism; barriers to treatment; and participants’ needs and priorities for improving GI health care. All interviews were conducted via Zoom and ranged from 22–110 minutes (mean = 35 mins). Participants could respond verbally or through the chat function, though none used chat exclusively. Accommodations included closed captioning, allowing a parent or support person to be present, and providing the list of main interview questions in advance. Four interviewers conducted the interviews: two autistic, one neurodivergent, and one neurotypical. Participants were given the option to choose whether they preferred to be interviewed by an autistic researcher.

### Data analysis

All participants gave prior permission for the interviews to be recorded, and the recordings were subsequently transcribed professionally by Rev. The analysis team consisted of four interviewers (CH, MF, KA, KD) and one community advisory board researcher with autism (FD), with additional analytic support from EP. We conducted a reflexive thematic analysis following Braun and Clarke,^12–14^ using an interpretivist-constructivist epistemological stance^15^ that views meaning as co-constructed between participants and researchers rather than discovered as objective fact. Coding and theme development were inductive and data-driven, allowing participants’ narratives and language to guide analytic direction rather than a predetermined theoretical framework. All five coders independently reviewed transcripts, noted potential codes and themes, documented analytic impressions, and engaged in iterative discussion to shape the codebook and emerging thematic structure. Before coding was completed, the team convened to discuss preliminary findings. Throughout the process the analysis team along with EP met regularly to refine codes, address coding questions, and finalize the set of themes. Once the codebook and thematic structure were finalized through collaborative discussion, one researcher (KA) applied the agreed-upon codebook to all transcripts to conduct a systematic analysis.

Our analytic process was informed by reflexivity^16^, acknowledging that our interpretations were shaped by the diverse neurotypes, lived experiences, and disciplinary training represented on the research team. Rather than seeking coder consensus or reliability metrics, we treated divergence as a source of insight and used discussion to deepen conceptual clarity. Themes were refined collaboratively through repeated engagement with the data, analytic memoing, and discussion of areas of interpretive uncertainty until shared meaning and coherent thematic structure were achieved.

## Results

### Themes

#### Theme 1: “I generally end up just feeling like garbage all the time.”

Participants repeatedly emphasized the enormous toll that their GI symptoms have on their lives. Managing symptoms requires an exhausting outlay of energy and affects every aspect of life, interfering with functioning and often strongly curtailing opportunities.

##### Subtheme 1.1: “it would be pretty frequent that I wouldn’t feel good”

Across interviews, participants emphasized that GI symptoms were not isolated medical events but ongoing disruptions that shaped multiple dimensions of daily life. Many described physically taxing symptoms that were difficult to control or anticipate. As one autistic adult explained, *“I was skin and bones and I couldn’t eat…I felt like passing out constantly…I couldn’t function.”* Another participant explained that “*at this point, we use MiraLax to try to treat that. That at times has been a little too effective*,” reflecting how even recommended treatments could cause new problems. Others described more severe episodes, including breathing interference (“*the GERD aspects of things, it sometimes interferes with her breathing*”) and emergency care (“*we were in the emergency room until four o’clock in the morning*”), illustrating how participants experienced symptom severity as escalating into acute medical crises. The cumulative toll of this ongoing management was evident as a participant shared, “*I was barely awake but waiting for the pharmacy to open*.” These ongoing physical disruptions were closely tied to emotional experiences, shaping how participants understood and related to their bodies.

##### Subtheme 1.2: “it’s almost like it turns in on itself and then your gut just hates you”

Beyond physical symptoms, participants described a profound emotional strain, including fear, embarrassment, and frustration. Concerns about public accidents were common and deeply distressing; as one participant shared, “*I’m really scared that I’m going to have an accident*,” while another described the emotional fallout as “*it’s humiliating*.” Some reflected on a sense of bodily betrayal, describing that *“it makes me feel like my body hates me*.”

##### Subtheme 1.3: “What happens if I have to pull off to the side of the road and explode?”

GI symptoms also had a direct and limiting effect on daily routines, movement, and autonomy, often requiring families to organize life around bowel movements and access to bathrooms. One caregiver shared, “*we don’t go anywhere until she has a bowel movement.*” An autistic adult noted, “*I had an exact routine, so I knew what to expect. I knew what my bowels were going to do, I knew what my brain was going to do. I knew what everything was going to do.*” Despite extensive planning and preparation to manage daily risk, additional layers of contingency planning were still required for this same participant to be able to drive to work in the morning, “*I have a 32-ounce cup in my car for emergencies, and it has happened.*” On days when treatments created excessive urgency, many times participation was simply not possible: “*if the MiraLax is being too effective, then we can’t do anything that day*.” Managing symptoms often involved extensive tracking and monitoring, with one participant stating, “*It’s definitely mentally and emotionally draining…that planning ahead of ‘Okay, if I eat this thing, am I going to experience this pain later? What are my plans for later on,’ and planning around that.*”

##### Subtheme 1.4: “Will I ever be able to date or do anything?”

Participants described social and relational consequences, noting that GI symptoms could shape comfort, privacy, and participation in shared environments. For some, even identifying an acceptable restroom was challenging, as in “*she’s got some challenges around what bathroom she’s comfortable using*.” Others described scaling back travel (“*we don’t take trips anymore*”) or opting out of eating with others (“*I don’t eat when I’m out with people*”), signaling that GI symptoms directly influenced opportunities for connection, leisure, and community engagement. Another participant reflected on how their GI symptoms have impacted opportunities for romantic connection, saying, *“It’s probably one of the most embarrassing things that I have to deal with, especially in dating, is I tend to get sick on my first date or meeting with people. Back when I was more involved in the dating world, I would nearly feel like I’m about to defecate myself. And it’s very embarrassing because I don’t feel like I can control it.”*

#### Theme 2: “that was my most recent trigger”

Participants described a wide range of potential triggers that appeared to influence the onset or severity of GI symptoms including stress, overstimulation, alcohol or specific food items, experiencing a lack of control, medications, interruptions to routine, menstruation, exercise, and “*eating in general*”. Across accounts, participants framed potential triggers as dynamic, interacting, and often ambiguous, summarizing the experience as: *“…is it my gut? Is it my stomach? Is it my digestion? Is it stress? I don’t know.”*

##### Subtheme 2.1: “but then sometimes I feel like the opposite would happen”

On the whole, predictability of GI symptoms and triggers was elusive. Participants emphasized difficulty identifying consistent causal patterns over time, noting that reactions fluctuated unexpectedly across days and circumstances. One participant explained, *“I can’t actually tell you definitively of when they started, how frequently they changed or how frequently they come on. Sometimes literally I’ll have like 3 good days in a row. And then sometimes I could just be dying on this day…”* For many, this lack of reliable or repeatable patterns made it challenging to establish clear management strategies or anticipate symptom escalation.

A number of participants described food-related factors, though not always in straightforward or linear ways. Eating, not eating, and the type or amount of food consumed were all mentioned as possible contributors, sometimes in contradictory ways. One participant reflected, *“Eating is more linked to pain in my stomach and so I try not to eat…”* while another noted that reactions did not align with expected patterns, *“You’re having these inconsistent reactions to food…it doesn’t make sense why…”*

##### Subtheme 2.2: “It depends on what life events are going on.”

Beyond food-related triggers, participants emphasized the importance of contextual factors in shaping the onset and severity of GI symptoms, including emotional, situational, and physiological contexts. Several participants described stress and heightened emotions as linked to symptom exacerbation. One participant reflected, “*It was from stress, because I was selective mute. That always just means pressure to speak…Life was just so extremely horrible…my only memories of doing anything are when I felt like I was going to vomit,*” illustrating how emotional overwhelm was experienced as directly embodied through GI distress.

Participants also highlighted situational factors, particularly disruptions to routine, as influential. Maintaining consistency was described as critical, with one participant noting, “*…if I deviate from my normal day-to-day schedule on a weekend, it’s extremely difficult to impossible to produce a bowel movement for that day.”* Others pointed to physiological cycles as part of this contextual landscape, including hormonal changes: “*Oh, and then periods also. I don’t know if that’s something that triggers it, but when it gets really bad…I will drink magnesium citrate and/or get an enema*.” Together, these accounts reflect how participants experienced GI symptoms as deeply context-dependent, shaped by the interaction of internal states, situational demands, and bodily rhythms.

#### Theme 3: “I have a love/hate relationship with my health system right now.”

Participants described healthcare as both essential and deeply taxing, characterizing their relationships with health systems and providers as emotionally demanding and shaped by negative past experiences. While care was often sought in response to worsening GI symptoms, many participants described a paradoxical pattern in which the process of seeking care itself intensified stress and, in turn, potentially exacerbated symptoms. As a result, healthcare engagement was experienced not as a neutral intervention but as part of a recurring cycle in which care-seeking created both emotional and psycho-physiological strain.

Participants emphasized the emotional labor required to prepare for, navigate, and recover from healthcare encounters, including managing uncertainty, advocating for themselves or their child, and revisiting prior distressing experiences. For some, this anticipatory stress caused symptom flare-ups, contributing to apprehension about seeking care even when it was clearly needed. Parents of autistic individuals described additional challenges as their children transitioned out of pediatric systems, noting new barriers to access, reduced provider familiarity with autism, and increased responsibility placed on families to coordinate and justify care. Together, these experiences illustrate how healthcare was not simply experienced as a source of support, but as a complex and sometimes destabilizing context that both responded to and intensified GI-related distress while often not providing meaningful answers or direction.

##### Subtheme 3.1: “the doctor just kept dismissing that”

Several participants felt their concerns were not taken seriously or that symptoms were minimized, leading some individuals to question the usefulness of seeking care. Some felt this dismissal was explicitly because they are autistic.

As one participant put it, *“Yeah, they weren’t concerned at all.”* Another stated, *“I have yet to find a provider that has any idea how to help us.”,* while an autistic adult explained, *“I’m having to actively find a doctor that is going to think outside the box because I’ve been dismissed as anxiety or drug-seeking or something else.”* A parent of an autistic adult child adds, *“At some point, I just became defeated by the fact that you’d go to a doctor and they were no use at all. They weren’t helpful and it becomes, well what’s the point of continuing to show up to a gastroenterologist and talk about these things when they don’t have any answers, they don’t have any solutions.”*

For several participants, these experiences connected to their autism diagnosis and identity, influencing how they interpreted past and present interactions. One person reflected, *“And then, finding out that I’m autistic in 2020, I remember just thinking about experiences before being diagnosed, and then it made sense why things were so invalidating anyway and I didn’t get a lot of answers.”* Another participant described how diagnosis reshaped their perspective on navigating care, stating, *“Like I said, when I realized I was autistic a couple years ago, everything started to make sense in terms of how I approach healthcare.”* On the flip side, others commented on how their diagnosis can affect how their healthcare providers interpret an interaction, *“It makes me wish that I knew a better way to advocate for myself because when I’m in those situations where I’m having to talk to them, sometimes I’m too nervous that a lot of things go out the window. I’ll literally forget why I’m there sometimes. And I don’t like it because to me, I feel like that opens the door to be infantilized and then dismissed and I can’t have that.”*

##### Subtheme 3.2: “it’s like a maze…it was close to causing a meltdown for me**”**

Participants pointed to system-level challenges affecting access and coordination, with some describing confusion or burden related to navigating referrals, primary care requirements, or organizational structures.

Challenges common to and characteristic of many autistic people further exacerbate this already complex and challenging process. One participant shared, *“It’s all boiled down to, ‘You need to get a [primary care provider].’ It’s all in the same system and I know that, but I just know what I’m going to have to go into…I feel like there’s just barriers and it’s just hard.”*

Participants described emotional, cognitive, or organizational effort required to communicate effectively with providers, sharing, *“I come in there with all of my binders and documents, and they just tell me what I already know*.” A parent of an autistic adult child with cognitive impairment commented, *“And at this point we’re starting all over again with needing to try to find specialists and providers. And now I need to find people in the adult world. And that I’m running into problems that in spite of being her guardian, depending on who the provider is, they lose their ever loving mind and are really mean when I’m trying to reach out and advocate and to get a conversation going with someone, or really it’s going to be my words that they’re going to rely on. She can talk but she’s not going to be super helpful. And so I just feel really stuck because I don’t know how to make any of it better…”*

Executive functioning challenges add to the burden, *“And keeping appointment times is just generally hard for me…That cuts across specialties. The only appointments I consistently show up to are my psychiatry appointments. I even blew off the ENT, even though my…I still can’t hear. And I think it would be helpful to me if there was a way to reschedule my appointments online or something because picking up the phone and making a phone call is kind of difficult for me. Also, just the confusion. Because I moved here from LA a year ago and I had totally different doctors at [hospital], and navigating the [hospital] system was totally different because I had a different kind of health insurance plan.”*

Literal thinking can pose additional communication challenges*, “I have no idea why they ask me certain things. Are you married, is a question that a lot of doctors ask, but I always thought it was benign. I didn’t realize that what they’re really asking me is is there a chance you’re pregnant? They assume because you’re married, you’re having sex, and that’s wrong. They never asked me that. What they ask was, are you married?”*

##### Subtheme 3.3: “Self-advocating for that has been, at this point, traumatic”

Healthcare encounters were described as emotionally fraught and frequently traumatizing, shaped by prior negative experiences and uncertainty about how providers would respond.

Many participants described avoiding care due to prior traumatic experiences. One participant described it as, *“But in the moment, all of that, there’s some trauma right there that you just introduced for your patient, and now they’re scared. And maybe they don’t want to ask for help if this happens again,”* and another commented, *“I haven’t really gotten any other treatment since then, mostly because I was like, ‘I don’t want to deal with this. I want to avoid it.’*”

This can lead to a cycle of anxiety related to seeking care, further complicating the process of seeking care and managing symptoms, *“Like I said, the stress and anxiety…that’s also so common and tends to be connected to healthcare for me, and that contributes to the GI issues, which contributes to the stress, and so on.”* Another commented, *“I guess still though, every doctor’s appointment is a little nerve wracking because I know that there’s going to be some issue just because that’s how they are,”* highlighting anticipatory stress before appointments even began.

#### Theme 4: “So it’s just been a process of elimination and just constantly going to see the doctor.”

Participants’ negative experiences with healthcare professionals, combined with the unpredictability of their GI symptoms, meant that they often needed to adopt a range of approaches to manage their symptoms; reflecting ongoing experimentation, adaptation, and efforts to reduce discomfort or prevent escalation. These strategies varied widely, including dietary adjustments, pacing and portion control, medication or supplement use, environmental planning, sensory or emotional regulation, and acceptance-based responses.

##### Subtheme 4.1: “the most helpful thing is to avoid them”

Many participants framed management as minimizing aggravation, often through pacing, reducing physical or emotional strain, keeping to routine and structured environments, and avoiding triggers. Across accounts, management was described as practical, adaptive, and personalized, guided by perceived effectiveness, bodily awareness, safety, and the desire to minimize distress rather than fully eliminate symptoms.

One person described intentionally stepping back rather than intervening, explaining, *“I try to just let it be what it is. I don’t really try to manage it or anything because then I get more frustrated and end up making it worse.”* Another said, “*Yeah, so it’s just avoiding things. And I mean, I still end up having issues with it from time to time, but it’s not constant like it was before.”*

Others used proactive behavior adjustments, such as modifying diet or intake, with one participant sharing, *“I try to eat smaller portions, because if I eat more than what feels okay, I’ll have symptoms.”* For some, healthy eating was an ongoing aspiration rather than a consistent practice, as reflected in, *“I mean, I try to eat well, but I’m not always the best at that.”*

Understanding the sources of GI issues was particularly powerful in helping participants avoid triggers to manage symptoms, *“One of the biggest things that have helped me with my symptoms is knowing that I have Celiac disease…knowing those triggers have been super helpful.”*

Many participants described environmental planning and preparation, prioritizing spaces where they felt safe, comfortable, and able to manage symptoms privately and on their own timing. One participant explained, *“It’s easier for me to be home at my own toilet than at the work toilet, so if I’m not feeling good, I’d rather be home.”* Another explained, *“I’m always prepared for things. If I leave the house, is there something I might need that I never need, but I’m going to take with me anyway? I have all the supplies in your car that I’ll ever need”.* Others incorporated physical positioning to reduce discomfort, such as *“For reflux I sleep propped up, because lying flat makes it worse.”*

Autistic adults and caregivers described actively structuring daily life and incorporating sensory or emotional regulation practices as ways of minimizing uncertainty and stress, which they experienced as closely linked to GI symptom severity. Consistency and predictability were frequently emphasized as protective, particularly in the context of symptoms that were severe or difficult to anticipate. One caregiver explained, “*He’s pretty routine oriented*,” while another reflected on the positive impact of structured living supports, noting, “*We don’t know if it’s the diet, the routine, or the structure, but since he’s been living in supported living…it has helped*.”

Participants described sensory and emotional regulation strategies aimed at maintaining calm to reduce symptom intensity. Calming activities were experienced as indirectly influencing physical symptoms, as one participant shared, “*Sometimes there’s one particular piece of music that if I listen to it, it will help calm me down*.” Others emphasized intentional stress reduction more broadly, noting, “*I try to eliminate stress, because stress makes everything worse*.” Together, these accounts illustrate how participants sought to stabilize both their internal states and external environments in order to limit symptom exacerbation, reflecting an ongoing effort to manage GI symptoms by reducing unpredictability and emotional strain.

##### Subtheme 4.2: “it helps alleviate some of the GI symptoms, but still…”

Participants also discussed many dietary, pharmacological, and supplement-based strategies, though these were rarely framed as complete solutions.

*“First would be water, just drinking a bunch of water, making sure that I’m fully hydrated. And my diet is whole food plant based.”* One person described, *“But certain things, like just taking Protonix to help with reflux, that’s helpful.”*

A caregiver shared, *“When he is getting a full dose of [supplements], then his stools are soft and he goes every day without having to drink any milk substitutes.”* Another participant described supplement use geared toward inflammation, noting, *“He has a fairly limited diet…but we found some oils that help with inflammation.”*

Others discussed solutions aimed at managing their gut microbiome, *“My naturopath in particular really thought that it would be beneficial for me to actually do the GI stool test, where they test for all the bacteria, all the viruses that could be in my gut.”* This approach worked better for some than for others, *“honestly, the probiotics have been the biggest help for me,”* and, *“I have been taking a probiotic every night before bed*.

*That seems to have helped some, just not significantly.”*

##### Subtheme 4.3: “I have no idea what’s going on.”

While many participants found relief in avoiding triggers or actively managing their diets, exercise, sleep, and other modifiable elements of their lives, many have found nothing helps or that their efforts end in inconsistent results, perpetuating their puzzlement and taking an emotional toll, which itself often then requires additional efforts to manage their wellbeing.

*”I really have no idea, and I know I’m not alone, which I alluded to earlier is a big part of why I’m participating [in this study].”*

One participant discussed their doctors as being as puzzled as they are, “*Why do I keep having these sores in my mouth? I don’t know. It’s a real odd one. Everyone that I’ve talked to has been like, ‘I have no idea what that is. I’ve never heard of that,’ the doctors.”*

Even knowing what symptoms to describe to a healthcare provider can be fraught. “*A lot of times a doctor will say, ‘Well, you didn’t mention headaches.’ How would I know that was correlated with this? How in the world? It’s like knowing that bloating has something to do with ovarian cancer. Nobody knows that, and especially not autistic people who have never been talked to about their bodies or people who’ve never been talked to about things wrong with them.*”

##### Subtheme 4.4: “Yeah. So there is unity and the support in it, even if it’s because of something that’s maybe not your top choice.”

Whether or not participants have found solutions that work for them, social connection and support was identified as critically important to their wellbeing, their ability to navigate life and manage symptoms and GI events, and in remaining hopeful enough to persist in finding solutions.

The perpetual lack of solutions is often frustrating and demoralizing, *“I myself have had a hard time coping with it because I feel like I’m boxing shadows.”* However, being connected to the right people seems to help counter the effects of this frustration in multiple ways and can be key to eventually finding things that help. *“I have a therapist who’s absolutely wonderful, who helps me find the people that I need or finds people who can find the people that I need. He has a lot of great colleagues and knowledge about a variety of topics. So, he’ll often act as a sort of intermediary. Also, my mom is very helpful. She’s wonderful, and she will often find or help me find a good doctor.”*

Being able to talk openly with others was described as helpful in the workplace and at school, as others were then able to understand how to accommodate and support the participant in their day-to-day work and understand why that is important. *“Yeah. I would say definitely people that are also fighting that trend of just keep to yourself, don’t complain about anything. And it’s not so much a complaint. This is important information. Fighting through not feeling well, and I don’t mean just, ‘Oh, I just feel a little under the weather.’ But just constant pain, it doesn’t do anybody any good.”* One autistic student commented, *“Well, before my classes even start with the social disability services at my college, so I’ll send them a disability center notification, just letting them know about my Crohn’s disorder and my ADD and all that. So they know about it going into it.”*

Some relied on their connection to the autistic community, “*I feel like this community is really big on…We’re here for the self-help because nobody else has the tools yet. So we’re over here and making the tools ourselves. So I feel community wise, it’s great*.

*We’re putting through the effort and the work*,” giving the following advice to others, *“I really enjoy social media communities for autistic people. Because I find it’s super helpful. So for me, I think it’s, don’t have a fear of trying to talk to people on Twitter.”* Others relied on friends or family members to help support them as they continue to struggle without answers. *“I talk to my brother a lot about it because he also has very similar GI issues, we commiserate about it.”*

Supporters would frequently encourage participants to seek care rather than just live with a problem, “*Now it’s my best friend who’s in New York I’ll basically tell everything to. She’s the one that basically tells me when it’s so bad, convinces me to go to the doctor or whatever. Then, now my boyfriend I’ll tell some of it, too. Not all of it yet. But, usually my best friend. She’s 10 years older, so she is wise. I feel like she’ll tell me. She also has GI issues as well, so she kind of understands. I’m just comfortable enough with her and she’s comfortable with me that we’ll share things with each other that other people would not understand or that it would be inappropriate to talk to, I guess.”*

Other times, supporters would even proactively share the burden of managing their autistic friend’s GI issues, *“It’s really sweet. Some of my friends, when they find out that I have GI issues, they’ll make it a mission, know that the place that we’re going has food that I can eat. Or, if they’re making some sort of food, they’ll make sure that they don’t put milk or bread in it.”*

## Discussion

This qualitative study provides new insight into how autistic adults and caregivers experience and manage GI symptoms in daily life. Participants described symptoms as chronic, unpredictable, and highly impactful on physical, emotional, and social functioning, extending prior quantitative research focused on pediatric prevalence patterns.^4,17,18^ Our findings demonstrate that GI concerns persist into adulthood with substantial effects on autistic people’s wellbeing, including their autonomy, identity, and involvement in activities of life.

Participants’ narratives emphasized difficulties identifying consistent triggers and reliance on individualized trial-and-error approaches to symptom management. This aligns with chronic illness frameworks describing uncertainty, adaptation, and embodied expertise,^19,20^ but also suggests autism-specific pathways, such as sensory sensitivities, emotional regulation differences, and need for routine, which shape how symptoms are experienced, expressed, and interpreted.

Healthcare experiences described in this study reflect well-documented inequities faced by autistic adults, including dismissal of concerns, diagnostic overshadowing, communication barriers, and limited sensory accommodations.²¹^-^²³ However, our findings extend this literature by illustrating how these inequities interact directly with GI symptom trajectories and care seeking behaviors. Participants frequently described avoiding or delaying care following prior experiences of invalidation, contributing to prolonged distress, unmanaged symptoms, and escalating care concerns.

These dynamics can be understood through the lens of the triple-empathy problem, which emphasizes reciprocal breakdowns in understanding between autistic individuals, caregivers, and healthcare providers operating under differing communicative norms and assumptions^25^. Within this context, participants’ accounts suggest that GI symptoms were not only medically challenging but also epistemically vulnerable—often rendered as indecipherable or insufficiently credible within clinical encounters. This reflects a form of epistemic injustice, wherein autistic individuals’ embodied knowledge of their own symptoms was frequently discounted or deprioritized, limiting opportunities for timely diagnosis and effective intervention.

At the same time, participants demonstrated considerable resourcefulness and adaptive expertise in managing symptoms, navigating fragmented systems, and advocating for care in the face of persistent barriers. These findings align with prior work characterizing autistic adults as active health advocates and problem-solvers rather than passive recipients of care.^23,2□^, Taken together, our results highlight how systemic failures in recognition and communication not only undermine trust in healthcare systems but may also exacerbate physical symptoms by increasing stress, uncertainty, and avoidance—further reinforcing the cyclical burden described across themes.

Collectively, our findings suggest that 1) clinically, providers may benefit from learning about neurodiversity-affirming, trauma-informed, and sensory-aware approaches that validate lived experience, incorporate collaborative problem-solving, and broaden assessment beyond biomedically oriented checklists, and 2) research priorities include development of adult-appropriate assessment tools that integrate sensory and psychosocial factors, examination of physiological-sensory-stress interactions, and expanded use of participatory research designs.

Specific practice- and systems-level recommendations, generated in partnership with our community-board and research participants, will be reported in a separate manuscript. Our findings align with findings in other similar research that has led to the development of tools such as the SPACE framework^27^ and the AASPIRE Autism Healthcare Accommodations Tool^28^ which aim to facilitate better healthcare interactions and outcomes for autistic people. Both were developed by identifying core needs common to most autistic people, agnostic of autism expression or care setting, and articulating differences from interactions with non-autistic patients that providers may experience and should be prepared for, with suggestions on how to engage meaningfully and respectfully with their autistic patients.

### Strengths and Limitations

This study’s strengths include its community-based participatory research design, inclusion of autistic researchers throughout investigational and analytic processes, and use of reflexive thematic analysis, which supports depth, transparency, and contextual interpretation.^11–13^ Accessibility-focused interview design features and the inclusion of both autistic adults and caregivers also strengthened data richness. Limitations include limited racial/ethnic and socioeconomic diversity and underrepresentation of minimally speaking and higher-support-needs autistic adults, reflecting persistent inequities in research accessibility. The relative absence of higher-support-needs adults is particularly salient given they may have unique challenges with communication, which may be a massive unexplored barrier to getting adequate or appropriate support. Future studies should incorporate multiple communication methods, targeted recruitment strategies, and additional supports to reach a broader range of autistic adults.

In conclusion, GI symptoms in autistic adults are chronic, consequential, and shaped by interacting biological, sensory, emotional, and contextual factors. Improving care will require multi-layered, neurodiversity-informed approaches that recognize autistic adults as central knowledge-holders and active partners in research and clinical decision-making.

## Funding and Acknowledgments

We extend our deepest gratitude to the members of our Community Board, whose expertise, lived experiences, and commitment guided every stage of this project. We also thank the autistic adults and parents who participated in this study for generously sharing their time, perspectives, and experiences. Finally, we gratefully acknowledge SPARK, a project of the Simons Foundation Autism Research Initiative (SFARI), for their support in participant recruitment through SPARK Research Match.

Lastly, we wish to thank our funders. This work was supported by Autism Intervention Research Network on Physical Health (AIR-P) (PI Holingue & Faucett) and the Autism Science Foundation (ASF) (PI Holingue). This project is supported by the Health Resources and Services Administration (HRSA) of the US Department of Health and Human Services (HHS) under award UT2MC39440, the Autism Intervention Research Network on Physical Health. The information, content, and/or conclusions are those of the author and should not be construed as the official position of, nor should any endorsements be inferred by HRSA, HHS, or the US Government.

## Author Contribution Statement

Calliope Holingue: Conceptualization, Methodology, Formal Analysis, Investigation, Resources, Data Curation, Writing – original draft, review & editing, Visualization, Project Administration, Funding Acquisition

Margaret Faucett: Conceptualization, Investigation, Formal analysis, Project Administration, Supervision, Resources, Funding acquisition, Writing – original draft, review & editing

Katherine Almendinger: Investigation, Data Curation, Formal analysis, Software, Visualization, Writing – original draft, review & editing

Faith DiJulia: Validation, Formal Analysis, Writing – review & editing Kelsie Daley: Investigation, Formal Analysis, Writing – review & editing

Zachary Williams: Conceptualization, Methodology, Writing – review & editing

Susan Brasher: Supervision, Writing – review & editing

Joseph J. Gallo: Supervision, Writing – review & editing

Elizabeth Pellicano: Supervision, Methodology, Validation, Formal Analysis, Writing – review & editing

## Data Availability

The participants of this study did not give written consent for their data to be shared publicly, so due to the sensitive nature of the research supporting data is not available.

## Notes

### Competing Interest Statement

The authors have declared no competing interest.

### Author Declarations

Institutional Review Board Approval was granted by Johns Hopkins University: JHM IRB 00316729.

## References

1. Doherty M, Neilson S, O’Sullivan J, et al. Barriers to healthcare and self-reported adverse outcomes for autistic adults: a cross-sectional study. BMJ open. 2022;12(2):e056904.

2. Khachadourian V, Mahjani B, Sandin S, et al. Comorbidities in autism spectrum disorder and their etiologies. Translational Psychiatry. 2023;13(1):71.

3. Leader G, Abberton C, Cunningham S, et al. Gastrointestinal Symptoms in Autism Spectrum Disorder: A Systematic Review. Nutrients. 2022;14(7):1471. 10.3390%2Fnu14071471

4. Holingue C, Newill C, Lee LC, Pasricha PJ, Daniele Fallin M. Gastrointestinal symptoms in autism spectrum disorder: A review of the literature on ascertainment and prevalence. Autism Research. 2018;11(1):24–36.

5. Buie T, Campbell DB, Fuchs III GJ, et al. Evaluation, diagnosis, and treatment of gastrointestinal disorders in individuals with ASDs: a consensus report. Pediatrics. 2010;125(Supplement_1):S1–S18.

6. McCormick JB, Hammer RR, Farrell RM, et al. Experiences of patients with chronic gastrointestinal conditions: in their own words. Health and quality of life outcomes. 2012;10(1):25.

7. Holingue C, Poku O, Pfeiffer D, Murray S, Fallin MD. Gastrointestinal concerns in children with autism spectrum disorder: A qualitative study of family experiences. Autism. 2022;26(7):1698–1711.

8. Denny E, Weckesser A. Quality not quantity: The value of qualitative research. Bjog. 2022;129(10):1799.

9. Black N. Why we need qualitative research. Journal of epidemiology and community health. 1994;48(5):425.

10. Renjith V, Yesodharan R, Noronha JA, Ladd E, George A. Qualitative methods in health care research. International journal of preventive medicine. 2021;12(1):20.

11. Fletcher-Watson S, Adams J, Brook K, et al. Making the future together: Shaping autism research through meaningful participation. Autism. 2019;23(4):943–953. 10.1177/1362361318786721

12. Braun V, Clarke V. Using thematic analysis in psychology. Qualitative research in psychology. 2006;3(2):77–101.

13. Braun V, Clarke V. One size fits all? What counts as quality practice in (reflexive) thematic analysis? Qualitative research in psychology. 2021;18(3):328–352.

14. Braun V, Clarke V. Reflecting on reflexive thematic analysis. Qualitative research in sport, exercise and health. 2019;11(4):589–597.

15. Creswell JW, Poth CN. Qualitative inquiry and research design: Choosing among five approaches. Sage publications; 2016.

16. Dodgson JE. Reflexivity in qualitative research. Journal of human lactation. 2019;35(2):220–222.

17. McElhanon BO, McCracken C, Karpen S, Sharp WG. Gastrointestinal symptoms in autism spectrum disorder: a meta-analysis. Pediatrics. 2014;133(5):872–883. 10.1542/peds.2013-3995

18. Madra M, Ringel R, Margolis KG. Gastrointestinal issues and autism spectrum disorder. Child and adolescent psychiatric clinics of North America. 2020;29(3):501–513. 10.1016/j.chc.2020.02.005

19. Bury M. Chronic illness as biographical disruption. Sociology of health & illness. 1982;4(2):167–182.

20. Charmaz K. Good days, bad days: The self in chronic illness and time. Rutgers University Press; 1991.

21. Kang LR, Barlott T, Turpin M, Urbanowicz A. A trial of the AASPIRE healthcare toolkit with Australian adults on the autism spectrum. Australian Journal of Primary Health. 2022;28(4):350–356.

22. Nicolaidis C, Raymaker D, McDonald K, et al. The development and evaluation of an online healthcare toolkit for autistic adults and their primary care providers. Journal of general internal medicine. 2016;31(10):1180–1189.

23. Nicolaidis C, Raymaker DM, Ashkenazy E, et al. “Respect the way I need to communicate with you”: Healthcare experiences of adults on the autism spectrum. Autism. 2015;19(7):824–831.

24. Raymaker DM, McDonald KE, Ashkenazy E, et al. Barriers to healthcare: Instrument development and comparison between autistic adults and adults with and without other disabilities. Autism. 2017;21(8):972–984.

25. Shaw, S. C., Carravallah, L., Johnson, M., O’Sullivan, J., Chown, N., Neilson, S., & Doherty, M. (2024). Barriers to healthcare and a ‘triple empathy problem’may lead to adverse outcomes for autistic adults: A qualitative study. Autism, 28(7), 1746–1757.)

26. Vaughn, L. M., & Jacquez, F. (2020). Participatory research methods–choice points in the research process. Journal of participatory research methods, 1(1).

27. Doherty, M., McCowan, S., & Shaw, S. C. (2023). Autistic SPACE: a novel framework for meeting the needs of autistic people in healthcare settings. British Journal of Hospital Medicine, 84(4), 1–9.)

28. Nicolaidis, C., Raymaker, D., McDonald, K., Kapp, S., Weiner, M., Ashkenazy, E.,…& Baggs, A. (2016). The development and evaluation of an online healthcare toolkit for autistic adults and their primary care providers. Journal of general internal medicine, 31(10), 1180–1189.)

